# Interprofessional Learning in Immediate Life Support training improves simulated patient related outcomes

**DOI:** 10.1101/2019.12.31.19016170

**Authors:** Jeremy Charles Morse, Craig William Brown

## Abstract

**Aim of the study:** To assess team performance in implementing time critical key interventions during a simulated resuscitation after participating in either an interprofessional-learning (IPL) or uniprofessional-learning (UPL) Immediate Life Support training course (ILS).

**Introduction:** Much of the published work on team-based simulation training has measured the lower levels of Kirkpatrick’s hierarchy of evidence and effectiveness. This study aimed to ascertain if interprofessional team training could improve a higher level of outcome such as behaviour and patient outcomes.

**Methods:** A retrospective quantitative analysis of time critical points in a simulated cardiac arrest resuscitation, from a previous randomised study on the effects of Interprofessional Learning in ILS. The video recordings from the original study consisting of medical (n=48) and nursing (n=48) students were analysed to mark when either the IPL or UPL team performed a time critical intervention.

**Results:** Five time-critical points for interventions were identified; confirmation of cardiac arrest, commencement of initial CPR, rhythm check, time to 1^st^ shock and delay in restarting CPR. Parametric testing of each of these time-based criteria were subjected to an independent sample *t-test* with statistically significant findings in three of the five criteria in favour of those who had undertaken the interprofessional learning.

**Conclusion:** Our results demonstrate that through an IPL approach in ILS there was a statistically different improvement in mean times to performing time-critical interventions, which if transferred to the clinical environment could improve and impact on both change of behaviour and patient outcomes in Kirkpatrick’s higher levels of evidence and effectiveness.

Though this study shows that team behaviour and performing time-critical interventions can improve in the short-term, we acknowledge that further longitudinal studies are required to ascertain the effect of such improvement over time. So that both as researchers and educators we can make healthcare teams work safer and more efficiently to improve patient outcomes.

What this paper adds

What is already known
Like simulation, intuitively we know that Interprofessional Learning and Team training should make a difference to clinical practice for those involved. The majority of research reported, is measured at the lower levels of Kirkpatrick’s hierarchy. This retrospective video analysis of a previous study aimed to look at the functioning between interprofessionally and uni-professionally trained teams in performing time-critical interventions in a simulated cardiac arrest.

What this study adds
Our study suggests that the use of Interprofessional Learning in resuscitation training does have an effect on performance during the management of a simulated cardiac arrest which could improve and impact on both change of behaviour and patient outcomes in Kirkpatrick’s higher levels of evidence and effectiveness.

## Introduction

Resuscitation training continues to be a core element of many undergraduate healthcare curricula. Standardised resuscitation courses, such as the Resuscitation Council UK’s Immediate Life Support (ILS) course^1^, are often structured to provide healthcare learners with an algorithmic approach to managing the critically unwell patient. Whilst these courses are delivered to learners from a number of disciplines at postgraduate level, often at an undergraduate healthcare level they are provided in a uniprofessional context. However, this is despite in practice almost always occurring as a multi-professional team-based response. Though studies of simulated cardiac arrest have been reported, the investigation of team-based factors in real life cardiac arrest scenarios remains challenging.^2^ Therefore, simulation-based studies such as this one, can offer educational researchers surrogate with markers in respect to measuring complex team-based indicators in a cardiac-arrest resuscitation.^3,4^

From the resuscitation literature it is known that in regards to patient outcome, for each minute of delay to defibrillation, when the patient is in a shockable cardiac rhythm, the probability of survival to discharge reduces by 10-12%.^5^ Therefore, the effective management of a cardiac arrest resuscitation requires a team-based approach both in training and in practice^4^. With previous studies showing that teams who were trained using an interprofessional ILS method, had improved overall team-working scores than those who had been trained uni-profesionally.^6,7,8^

Much of the published work on team-based simulation training has measured outcomes on the lower levels of Kirkpatrick’s hierarchy of evidence of effectiveness, i.e. learner outcomes such as self-reported views, perceptions or attitudes, rather than the process elements of quality care^9,10^.

This study, in line with the 2015 report from the Institute of Medicine, hopes to provide another step towards building more solid evidence in linking interprofessional learning to patient outcomes.^11^ It was therefore hypothesised that if interprofessional learning can improve team performance, then this could also be seen in time critical points throughout the simulated resuscitation, thus leading to potential better patient survival and provide potential evidence at Kirkpatrick levels 3 & 4 (‘behaviour’ and ‘patient outcomes’).

### Ethical Approval

Ethical approval for the original study was obtained from both the School of Nursing Ethical Review Panel, Robert Gordon University (RGU) and the College Ethics Review Board, College of Life Sciences and Medicine, University of Aberdeen (UoA) and included approval for limited disclosure to minimise any perceptual bias regarding Inter-Professional Learning (IPL). In this way all participants were blinded as to the study objectives regarding the effect on the team performance/interaction during a simulated resuscitation. The participants were however made aware that it was a study to evaluate the teaching of ILS to final year medical and nursing students.

## Methods

### Design

A retrospective quantitative analysis of time critical points in a simulated cardiac arrest resuscitation, from a previous randomised study on the effects of Interprofessional Learning in Immediate Life Support (ILS).^8^

Participants in the original study^8^, were randomised by student profession (medical n=48 and nursing n=48) and attended either an interprofessional or uniprofessional ILS course. Each course followed the UK Resuscitation Council one day programme including facilitated discussions, A to E assessment of the deteriorating patient, airway and safe defibrillation workshops and six cardiac arrests scenarios in which each candidate had an opportunity to team lead in a cardiac arrest. Throughout the course the participants were orientated to and used the Laerdal® Mega Code Kelly (advanced) mannequin along with all the equipment that they would require to perform a resuscitation. For the recorded scenario and to ensure familiarity the same resuscitation equipment and mannequin was placed in the simulation laboratory of the University of Aberdeen Medical School, Clinical Skills Centre. To ensure consistency for data analysis each team received the same cardiac arrest scenario, a patient presented in cardiac arrest and in a shockable rhythm (ventricular fibrillation) (*see appendix 1*). Each teams’ scenario was recorded using the SMOTS ™ fixed recording system (Scotia UK plc, Edinburgh UK) installed in the simulation rooms allowing for detailed analysis of each groups performance.

Following the initial ILS courses approximately two weeks post intervention participants from the respective cohorts were invited to the simulation laboratory to undertake a simulated resuscitation in randomly assigned teams comprising equal numbers of medical (n=3) and nursing (n=3) students based on their original ILS course with a total of sixteen teams in total (IPL n=8 and UPL n=8). In the original study the aim was to look at if the intervention of interprofessional ILS improved overall team performance in managing a simulated cardiac arrest as opposed to those who were trained uniprofessionally. On the day of the scenario this was completed by an experienced ALS instructor who provided feedback on the team’s performance immediately afterwards. While the video recording was evaluated and scored by a single independent reviewer (an experienced clinician and ALS instructor) who was blinded as to which course they had undertaken. The data from both the instructor on the day and the blinded reviewer were then entered in to the SPSS 10 statistical software and analysis undertaken.

In this retrospective quantitative analysis of the original study data it was proposed that the time taken to by each of the teams to complete five time-critical interventions be investigated to ascertain if there were any statistically significant differences between the provision of IPL or UPL ILS courses.

Following advice from the department of medical statistics at the University of Aberdeen it was decided to employ independent sample t-test to compare the time taken to complete key time-based interventions between the two cohorts, with a starting point from when the team entered the room. These key time-based criteria, measured in seconds were; confirming cardiac arrest, commencement of chest compressions, time to delivery of first shock and delay in restarting CPR post shock. The resultant qualitative data, from the retrospective analysis, were then imputed into the SPSS 10 statistical software and parametric testing applied.

## Results

Video recorded data for eight ‘IPL’ and eight ‘UPL’ trained teams, comprising of a total of ninety-six participants were available for the follow up cardiac arrest simulation scenario and each time-based criterion was subjected to an independent sample t-test are summarised and displayed in *table 1*

**Table 1 –.**
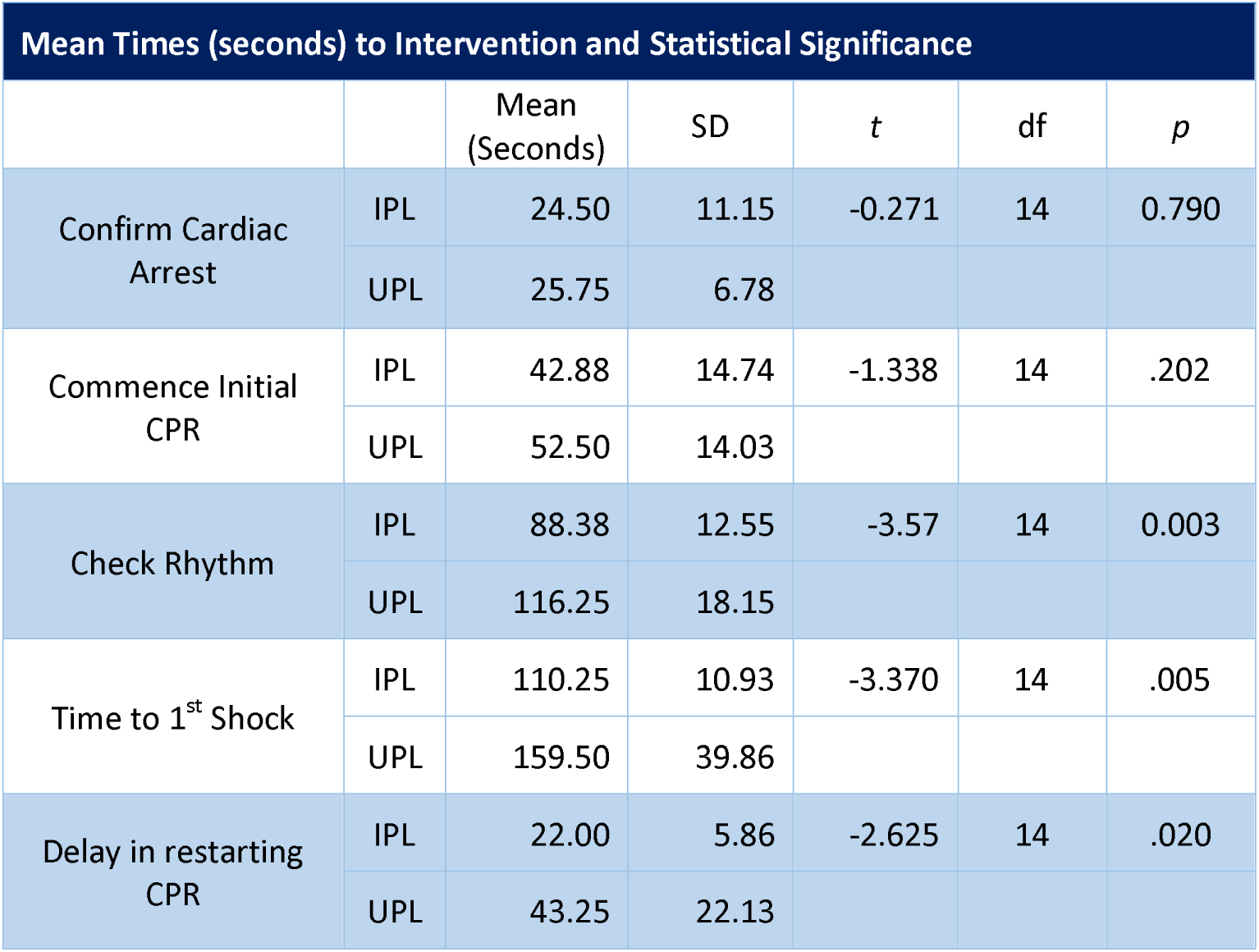
Mean Times in seconds from entering the room.

An independent-samples t-test was conducted to compare the time taken in seconds, to confirm cardiac arrest by the IPL and UPL teams which showed that there was no statistical significance in the time taken for UPL teams (M = 25.75, SD = 6.78) and IPL teams (M = 24.5, SD =11.15; t (-.271), p =0.790). (*Figure 1*). Similarly, in regards time to commencement of CPR an independent-samples t-test was conducted on the number of seconds taken by the IPL and UPL teams which again showed despite the IPL cohort commencing compressions in a mean time of 10 seconds faster than the UPL teams, there was no statistical significance in time taken between the UPL teams (M = 52.5, SD = 14.03) and IPL teams (M = 42.88, SD =14.74; t (-1.34.), p =0.202). (*Figure 2*).

**Figure 1 –.**
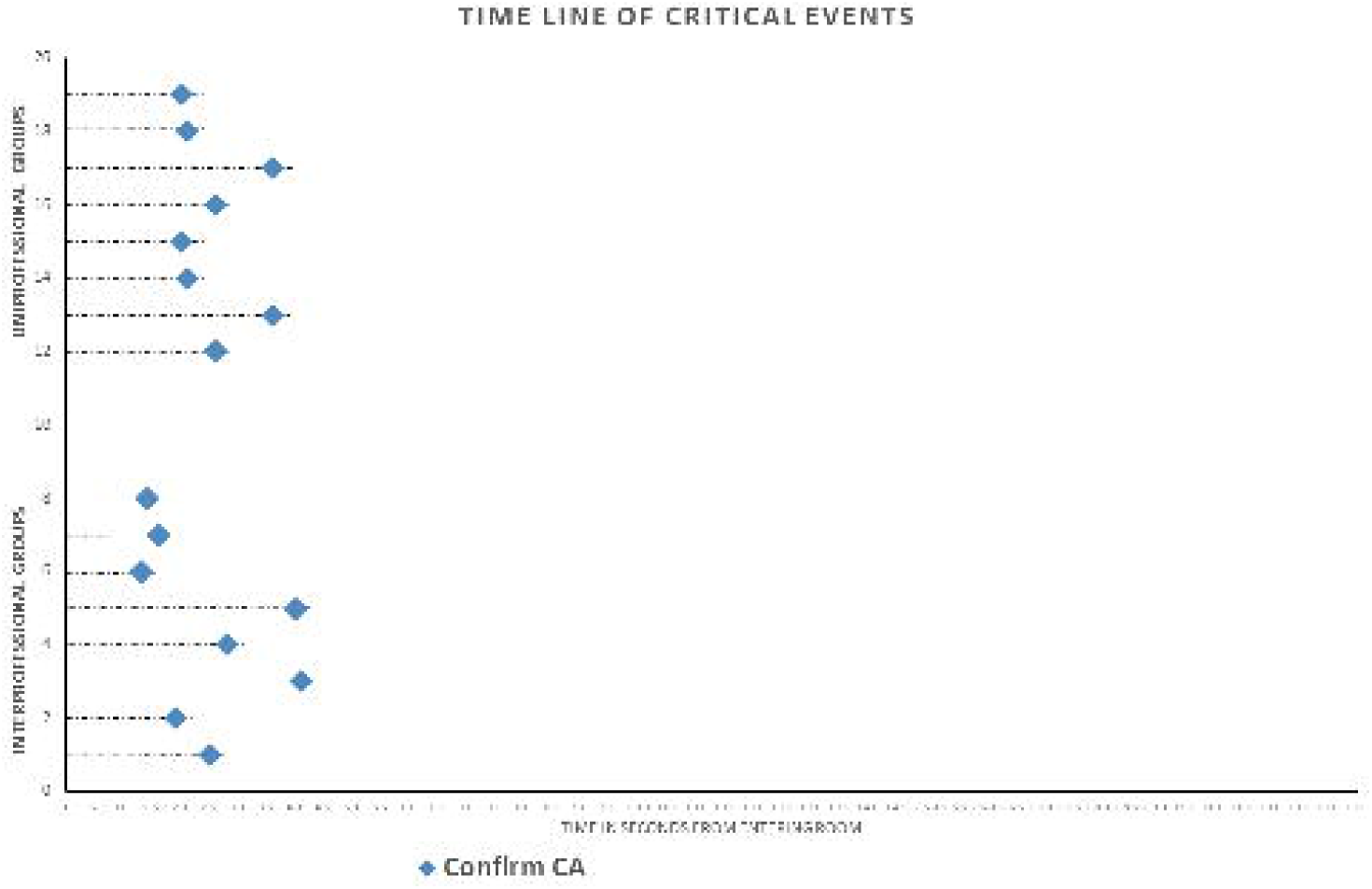
Time to confirming cardiac arrest.

**Figure 2 –.**
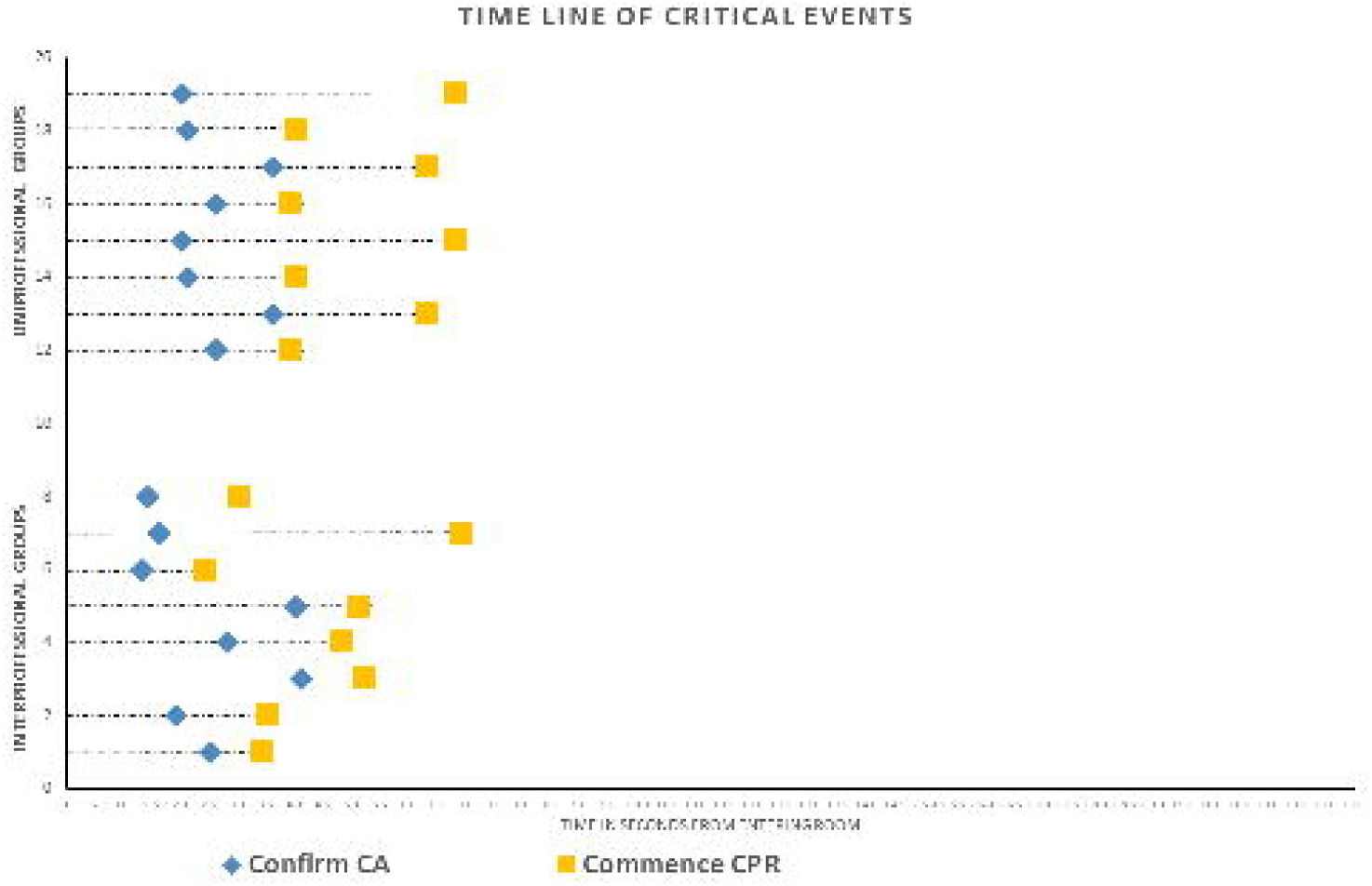
Time to commencement of CPR.

When analysing the differences between the time taken between the groups to perform the first ‘rhythm check’ (a process involving connecting the mannequin to the defibrillator whilst CPR was ongoing) an independent-samples t-test was conducted that showed statistical significance in time (seconds) taken between the UPL teams (M = 116.25, SD = 18.15) and IPL teams (M = 88.38, SD =12.55; t (-3.57.), p =0.003). (*Figure 3*). In regards the time critical point of delivering the ‘first shock’, the independent-samples t-test conducted also showed there was statistical significance in the time in seconds taken between the UPL teams (M = 159.50, SD = 39.86) and IPL teams (M = 110.25, SD =10.93; t (-3.37.), p =0.010). (*Figure 4*). The final key-time based criteria analysed was the recommencement of CPR after the first shock. The independent-samples t-test was conducted and once again showed that there was statistical significance in time taken between the UPL teams (M = 43.25, SD = 22.13) and IPL teams (M = 22.00, SD =5.86; t (-2.625.), p =0.030).

**Figure 3 –.**
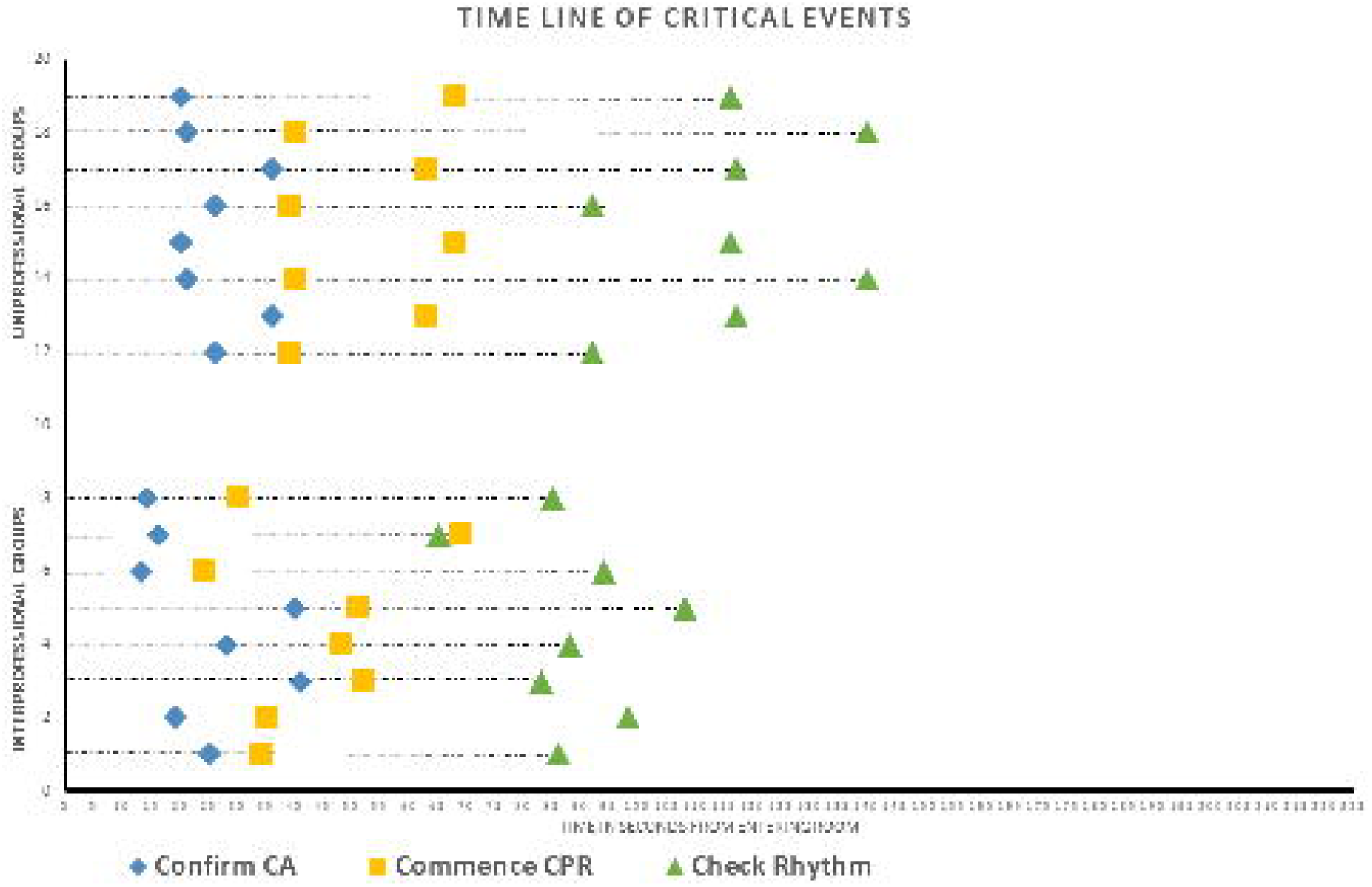
Time to check cardiac rhythm.

**Figure 4 –.**
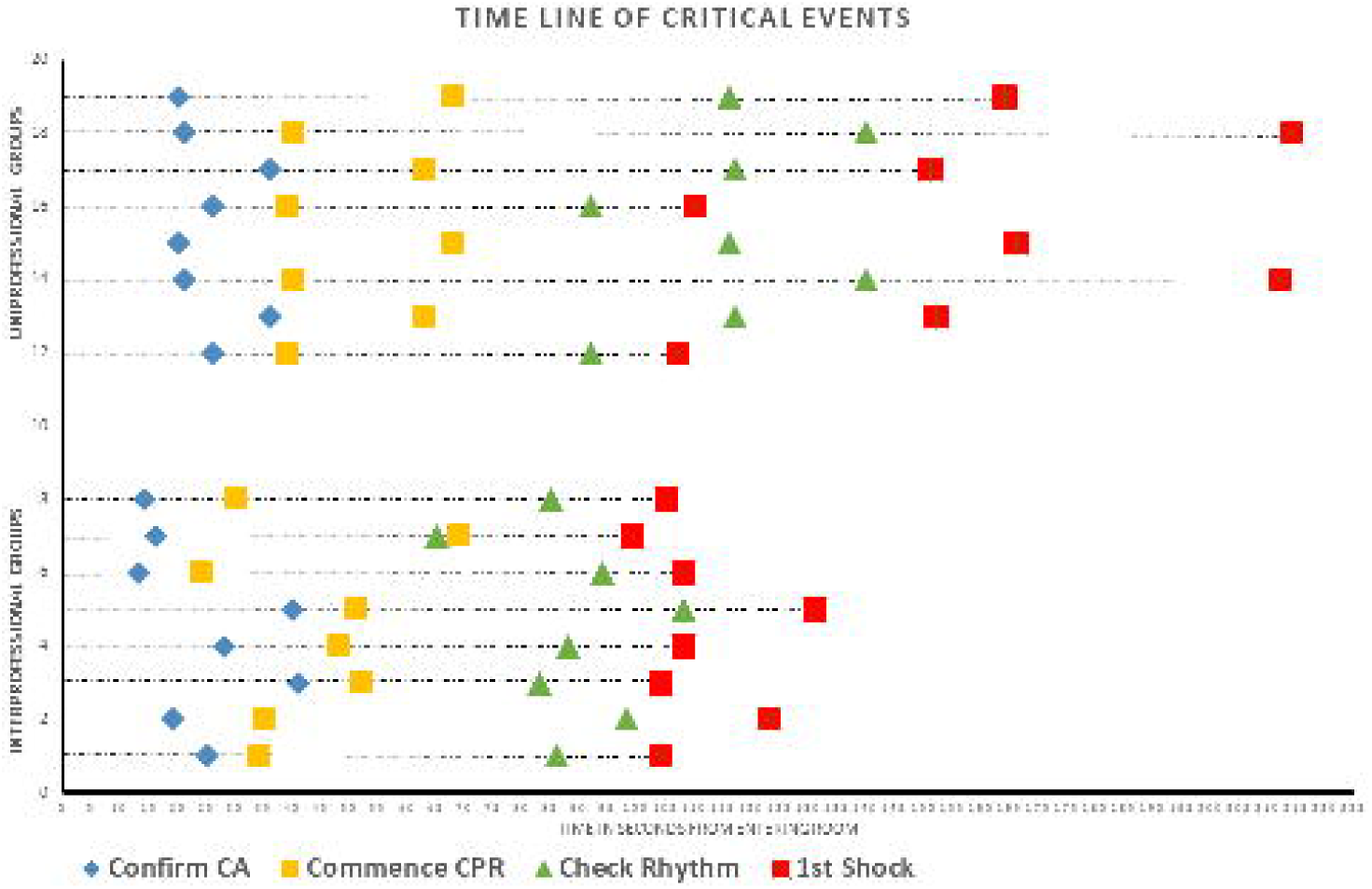
Time to delivering first shock.

## Discussion

It is recognised that interprofessional learning is essential in the training and education of healthcare professionals in order to equip them with the prerequisite skills, knowledge and attitudes to be able to work together in meeting the needs of patients.^12^ However, despite this there remains no clear path on how to assess the effective preparation of healthcare professionals to work collaboratively. It is therefore hoped that this study will assist in not only highlighting the positive impact that collaborative working can achieve, but also the impact on patient outcomes in a simulated environment, which can then be transferred to the clinical setting. ^13,14^ Within clinical practice most in-hospital cardiac arrests are managed by interprofessional teams, as opposed to uniprofessional groups or isolated responders, with an emphasis on the importance of good team work and leadership. Currently within the UK at undergraduate level, many of the mandatory life support courses continue to be delivered in a uniprofessional manner despite increased regulatory emphasis on team-based approaches to healthcare^15, 16^.

In the first component of this programme of research, it was identified that undergraduate students who were trained in an interprofessional format with ILS performed significantly better on TEAM scoring in a simulated cardiac arrest scenario^8^. Although the use of the TEAM score^17^ allowed the for the assessment of IPL versus UPL resuscitation team performance. This retrospective analysis has shown that IPL trained teams not only can achieve improved team performance but more importantly are able to perform better time-based outcomes in checking initial rhythm and most importantly a delivery of the ‘first shock’ in a simulated cardiac arrest than those trained uni-professionally. Whilst it can be argued that enhancing performance through interprofessional learning is a pre-curser to improving patient outcomes, it could also be suggested that this would impact on process related outcomes ultimately reflected in patient survival.

It has been well documented that in cardiac arrest situations, shorter times taken to complete time-critical outcomes were improved through standardised delivery of resuscitation training programmes such as the UK resuscitation Council ILS course.^1,12^ Indeed, it is the algorithmic teaching of such courses that encourage learners to promptly diagnose cardiac arrest, commence effective CPR and perform early defibrillation when indicated, that can maximise the chance of survival both in the pre-hospital and in-hospital situation. Furthermore, that in clinical practice, any delay in any of these key components or prolonged pauses in the algorithmic process are associated with poorer patient outcomes.^18,19^

From the analysis of this study’s data there are statistically significant findings that suggest interprofessional trained teams achieved the key time critical points of performing rhythm check and delivering the first shock faster than those who were trained uniprofessionally. Although it is acknowledged this remains a simulation-based resuscitation, these statistically significant results assist in progressing the related simulation-based education (SBE) literature on Kirkpatrick’s hierarchy of effectiveness at least to level 3 ‘behavioural change’ and potentially level 4 ‘impact on patient outcomes’.

To achieve this practising as a team and receiving feedback with the aim of improving overall performance has to be an integral component of interprofessional learning environment. This was the case in the delivery the initial ILS courses for both cohorts’, where all the participants would have received feedback and learning conversations upon their performance. Whereas with any feedback the aim is to identify any potential performance gaps and allow participants to reflect and reframe their thought processes and actions for future situations. Therefore, whilst those in the UPL cohort would have been given ways to improve on performance, it can be suggested that those in the IPL cohort benefited further from the feedback and learning conversations from a multidisciplinary team perspective and sharing the experience to work collaboratively in a resuscitation scenario. As a result, it was this feedback to the IPL teams, combined with the focused and deliberate and shared practise on the ILS algorithmic process that led to the statistically significant results identified in this study.

It has been reported that such deliberate practise in healthcare procedures leads to improved chronometry i.e. time taken to complete procedures.^20^ Whilst there was no data reported from either of the cohort’s initial resuscitation courses, the retrospective study data showed that those trained interprofessionally had significantly improved chronometry. Especially in relation to the delivery of first shock and recommencement of CPR, two of the key factors in the chain of survival and improved patient outcomes of patients in cardiac arrest.

Interprofessional learning as a pedagogical approach to training continues to rise as educators across the many professions embrace the concept that ultimately healthcare on the ‘shop floor’ occurs in teams. Further, that for IPL to be truly effective and have an effect on outcomes then all members should train together and be aware of each other’s roles, capabilities and ability to perform as part of a multidisciplinary team. It was strikingly evident during the video analysis that there was a notable difference in the way that the IPL trained cohort were aware of and respected each other whilst working together as a more cohesive team.

Although the impact of IPL on patient outcomes remains unclear, it is hoped that the evidence provided through this study will assist in furthering the knowledge base on which further research can continue to expand and promote the benefits of interprofessional learning.^21^

As SBE continues to be widely adopted and the drive for patient safety continues, interprofessional learning and deliberate practice such as in the case of resuscitation skills, allows for students to understand the equality of roles within the team. Indeed, the student participants in this study interacted and responded more positively following the combined profession training than those completing the uniprofessional training. Yet despite the growing evidence and the willingness from many educators to further utilise interprofessional learning, this is often still limited to just two professions rather than all those involved in a collaborative practice. A situation which, while challenging, may make such team-based practice less relevant.^22^ However, as has been demonstrated by the results of this study, the use of IPL between medical and nursing students significantly improved the overall team’s performance. Furthermore, that the use of IPL in resuscitation training allowed the team members to see the equality of role expectations, rather than the professional boundaries often assumed at an undergraduate level.

## Limitations to the study

It is acknowledged that analysing higher level Kirkpatrick related outcomes in randomized interprofessional simulation studies remain difficult, in part due to the total number of participants involved being divided into teams, in this case ninety-six participants forming sixteen teams. Also, that if there was a delay at the initial recognition of cardiac arrest then this would have a ‘knock on effect’ to the other key markers within the study parameters. However, the same could also occur in clinical practice and the statistically significant results of the study indicate that IPL has mitigated some of the argument.

## Conclusion

The initial study methodology of incorporating the TEAM tool to quantitively measure the performance of IPL and UPL trained medical and nursing students, indicated a statistically significant difference in team performance for those from the IPL cohort. This second and retrospective study of the data has further demonstrated that as well as increased team performance, those who underwent interprofessional learning completed the time critical key tasks of initial rhythm check and first defibrillation in a cardiac arrest faster than those who had the uni-professional training. Moreover, this study has identified and evaluated that as a direct result of interprofessional learning patient outcomes can be improved and adds to the evidence base that training in a multi-professional educational experience can have an impact not only on IPL but on collaborative care on patient outcomes and health both in the UK and worldwide.

Supporting recent recommendations,^23^ this study indicates that in undergraduate healthcare curricula and for which either basic or intermediate life support is required, such training should be done so wherever possible interprofessionally. Furthermore, that despite the low number of studies measuring the outcomes of behavioural change and benefits to patient outcomes, this research adds to the evidence that the use of IPL in resuscitation training can make a difference. To this end, further longitudinal studies are required to ascertain the effect of such improvement over time, so that both as researchers and educators we can make healthcare teams work safer and more efficiently to improve patient outcomes.

## Contributors

JM: participated in study design, data analysis and interpretation, drafting and revising the manuscript and approved the final version. CB: participated in study design, data interpretation, drafting and revising the manuscript and approved the final version.

## Data Availability

Data are available on reasonable request

## Acknowledgments

The Department of Medical Statistics for their continued support and advice

